# Knowledge, Attitudes, and Practices towards HPV Vaccination among Reproductive Age Women in a HIV Hotspot in the US

**DOI:** 10.1101/2022.09.12.22279885

**Authors:** Aasith Villavicencio, Gray Kelsey, Nicholas F Nogueira, Julia Zukerberg, Ana S Salazar, Lucila Hernandez, Patricia Raccamarich, Maria L Alcaide

## Abstract

**Background:** Human papillomavirus (HPV) is the most common STI in the US, responsible for cervical cancer and increased risk of HIV acquisition. Despite an effective HPV vaccine, women’s HPV vaccination coverage and rates remain far below desired levels. This study aimed to evaluate HPV knowledge, attitudes, and vaccination practices as well as factors associated with HPV vaccination among women of reproductive age living in Miami, Florida, a Southern US city with a high incidence of STIs and low HPV vaccination coverage.

**Methods:** From April to June 2022, 100 HIV-negative, cisgender, sexually active women aged 18-45 years were were recruited from the Miami community. Participants completed questionnaires using REDCap© electronic surveys validated questionnaires assessing socio-demographics and sexual behaviors; HPV knowledge, screening, vaccination practices; barriers and motivators to HPV vaccination. A cumulative HPV knowledge score (HPV score) was generated. Factors associated with HPV vaccination were analyzed by Chi-square, Fisher’s exact test, studentized t-test, and multivariate logistic regression (MLR).

**Results:** A total of 100 participants were enrolled, and 84 who knew their vaccination status were included in the analysis. Of these, 43 reported receiving at least 1 HPV vaccine dose (vaccinated group) and 41 reported never being vaccinated (unvaccinated group). Mean age was 24.7 (SD 4.03) years for the vaccinated group and 31.4 (SD 8.33) for the unvaccinated group. Mean HPV score was 18.9/29 (SD 6.05) for the vaccinated group and 9.1/29 (SD 8.82) for the unvaccinated group. Amongst vaccinated participants, 76.74% reported a history of HPV/Pap smear screening vs 87.80% in the unvaccinated group. Barriers to HPV vaccination included: 14.6% low-risk perception, 29.3% healthcare barriers, and 46.3% vaccine hesitancy and personal beliefs. Motivators t HPV vaccination included: risk perception and vaccine beliefs (71.42%), healthcare-related (60.71%) and social motivators (55.95%). In the first MLR, one-point increases in HPV score were significantly associated with higher odds of HPV vaccination until an HPV score of 16, and a one-year increase in age was associated with a 16% lower odds of HPV vaccination (aOR=0.84, 95% CI [0.72, 0.99]; p=0.035). Contraception use was also associated with HPV vaccination (aOR 8.36 (95% CI [1.41, 49.62]; p=0.020). Race, ethnicity, college education status, and number of sexual partners were not significant predictors of HPV vaccination. In the second MLR evaluating vaccination motivators as predictors of HPV vaccination, we found that individuals who were motivated by healthcare had 3.03 (95% CI [1.02, 9.00]; p = 0.046) times the odds of HPV vaccination compared to individuals without healthcare-related motivators.

**Conclusion:** Findings suggest suboptimal HPV knowledge and low vaccination rates among women of reproductive age. Public health efforts should focus on increasing basic HPV knowledge among women with little-to-no HPV knowledge to increase vaccine uptake.

## INTRODUCTION

In the United States, human papillomavirus (HPV) is the most common sexually transmitted infection (STI) among men and women.^1,2^ It has been estimated that yearly, up to 14 million individuals acquire a new HPV infection and 45,300 of those are diagnosed with HPV-associated cancers.^3,4^ While HPV vaccination has proven to be effective in decreasing the incidence of infection and HPV-attributable cancers, in 2019 only 47.0% of adults aged 18–26 years had received at least one dose of the HPV vaccine.^5^

Uneven HPV vaccination initiation and completion rates have been reported across states, by race and ethnicity, further widening disparities in HPV-related diseases.^6^ While some studies have suggested potential drivers for such disparities (e.g., health insurance, socioeconomic status, providers recommendations), results seem to vary depending on the population demographics.^7-11^ Hence, to reduce the gaps in HPV-related attitudes towards vaccination and to inform evidence-based tools that mitigate these disparities, a better understanding of the predictors of HPV vaccination is needed.

HPV knowledge has been used as a predictor of HPV vaccination acceptance and uptake.^12^ Thus, several states have committed funding to promote awareness of HPV and vaccination in the general population.^13,14^ Nonetheless, recent data suggest that HPV knowledge and general awareness have decreased overall in the U.S. since 2013.^15,16^ It is therefore important to assess the population-level awareness of HPV periodically to inform public health interventions aimed at promoting HPV awareness.

Compared to the national average, South Florida has a higher HPV-associated cancer incidence, lower HPV-vaccine completion rates, and a broad diversity in race/ethnicity.^2,17,18^ Particularly in Miami, prior reports have shown lower rates of HPV vaccine uptake and national cancer screening guidelines compliance -including cervical cancer-primarily among racial minorities and those with low socioeconomic status.^19^ Thus, this study aims at understanding the current HPV-knowledge, screening and vaccination practices among multiethnic women of reproductive age in Miami. Findings will shed light on identifying potential modifiable factors that can increase HPV vaccination uptake and reduce the incidence of HPV associated cancers.

## MATERIAL AND METHODS

### Ethical Considerations

The study was approved by the University of Miami (UM) Miller School of Medicine the Institutional Review Board (number: 20210504). Verbal consent was obtained from all participants before beginning the study assessment. A $15 dollar compensation was sent electronically or by mail to all participants who completed the survey.

### Study design and participants

This is a cross-sectional survey study conducted from April to June 2022. Participants were cisgender women, ages 18-45 years living in Miami, sexually active, HIV uninfected, and non pregnant. Participants were selected in a convenience sampling method through printed flyers, word of mouth, and participant registries from prior studies conducted by the research team at the Center for AIDS Research (CFAR) in Miami, Florida, including a study focused on understanding recurrence bacterial vaginosis and HIV risk among reproductive age women. Exclusion criteria included self-reported positive HIV diagnosis, history of abnormal pap smear or history of HPV diagnosis. Participants who were deemed eligible and provided consent were given the option to complete the study survey through a secure web-based system (self-administered via a unique web link using the University of Miami REDCap©) or by telephone by trained interviewers and logged into REDCap©. Time commitment for completion of the survey was 15 to 30 minutes.

### Study Assessments

Study tools included questionnaires collecting basic participant information and validated surveys. Sociodemographic variables were recorded including participant age, race, ethnicity, employment status, living situation and monthly household income. Medical history was also collected including obstetric and gynecological history, sexual and reproductive health, and history of genital infections or sexually transmitted infections.

The adapted Sexual Risk Behavior Assessment for adults, an established tool to assess HIV risk behaviors, was used to evaluate number of sexual partners, condom use, frequency and date of most recent intercourse, and alcohol and drug use. This survey has been previously used to assess risk behaviors. ^20^

A cumulative HPV score was calculated using the HPV knowledge, HPV testing and HPV vaccination survey, validated by Waller.^21^ This is a 29-item measure survey with psychometric properties established by classical test theory and item response theory (16 items - HPV knowledge score, 6 items – HPV testing (screening) score, 7 items – HPV vaccination score). Cronbach’s alpha and test-retest reliability values were reported as 0.838 (n=648) and 0.79 (n=226) respectively. In addition, Barriers to the HPV vaccination among unvaccinated participants and motivators to vaccination were assessed using a survey by Steben et al.^22^

### Statistical Analysis

Descriptive statistics were calculated using frequencies and mean/median, and groups were stratified by HPV vaccination status. Group comparisons between women who received at least one dose of HPV vaccination and no HPV vaccination were conducted using chi-square, Fisher’s exact test, studentized t-test, and Kruskal-Wallis test. Barriers and motivators to HPV vaccination were collapsed into categories. Predictors of HPV vaccination were analyzed through two multivariable logistic regressions (MLR). Inclusion of covariates in the model were based on previous literature, and the number of independent variables was bounded by sample size restrictions. Individuals with missing values for outcome and explanatory variables were not included in the final model. Linearity and multicollinearity assumptions were assessed using Box-Tidwell test, condition indices (CNI) ≥ 30, and variance decomposition proportions (VDP) ≥ 0.8; and goodness-of-fit was examined using Hosmer-Lemeshow test. Violation of linearity assumption of HPV score and log odds was corrected with the inclusion of a higher order HPV score term. Multicollinearity violations were corrected with recentering of age and HPV score around their medians. MLR results were presented as adjusted odds ratios (aOR) and 95% confidence intervals (CI) in forest plots. An α = 0.05 was used to determine statistical significance and analyses were conducted using SAS 9.4.

## RESULTS

### Sociodemographic Characteristics and Sexual Behaviors of the Respondents

A total of 487 women were identified as potential participants and contacted. Out of 157 women screened, 141 deemed eligible with questionnaires distributed via email. One hundred questionnaires were recovered (recovery rate of 70.9%). Among the 100 respondents, 43 (43.0%) women reported at least one HPV vaccine dose (i.e., vaccinated group), 41 (41.0%) reported not receiving any dose (i.e., unvaccinated group), and 16 (16.0%) women did not know their vaccination status. The latter group was excluded from our analysis. Sociodemographic characteristics are presented in Table 1.

**Table 1.**
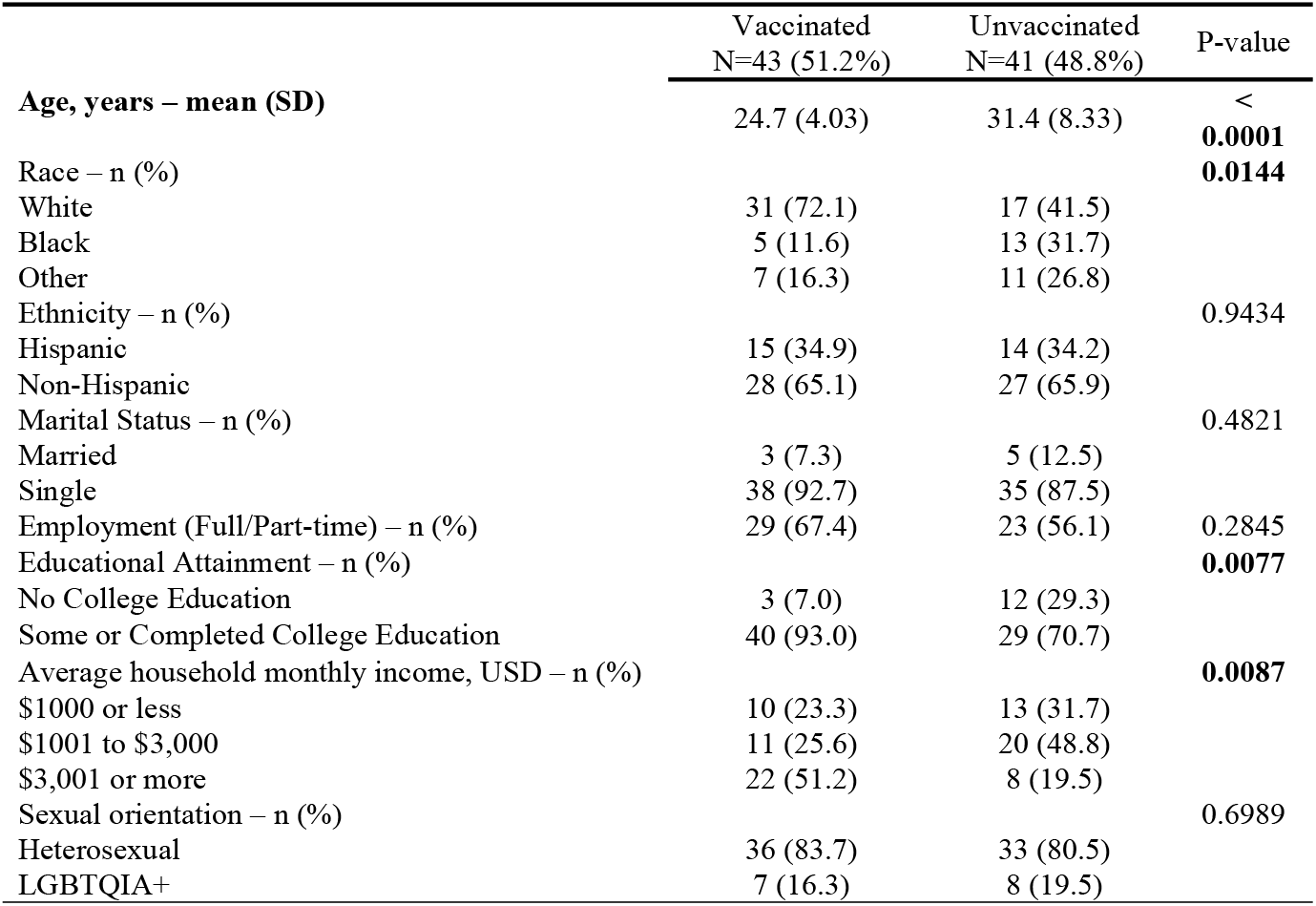
Sociodemographic Characteristics (N=84)

Table 2 reports the medical and gynecological history, sexual behaviors, HPV knowledge, screening, and vaccination practices. The vaccinated group was younger (24.7 vs 31.4 years, p < 0.0001), predominantly white (72.1% vs 31.5%, p = 0.0144), with some college education (93.0% vs 70.7%, p = 0.0077), higher average household income (51.2% vs 19.5%, p = 0.0087) compared to the unvaccinated group. Furthermore, vaccinated women had a higher proportion of current contraceptive use (69.8% vs 22.0%, p < 0.0001) and lifetime substance use (76.7% vs 39.0%, p = 0.0005); and lower proportion of lifetime pregnancy (9.3% vs 51.2%, p < 0.0001).

**Table 2.** Medical and Gynecological History, Sexual behaviors, HPV Education & Practices among study participants (N = 84)

|  | Vaccinated<br>N = 43 (51.2%) | Unvaccinated<br>N = 41 (48.8%) | P-value |
| --- | --- | --- | --- |
| **Medical and Gynecological History** |  |  |  |
| History of previous HIV testing - n (%) | 25 (58.1) | 25 (61.0) | 0.7912 |
| PrEP Aware - n (%) | 27 (62.8) | 25 (61.0) | 0.8640 |
| PrEP Use - n (%) | 0 (0.00) | 0 (0.00) | N/A |
| Current Contraception Use - n (%) | 30 (69.8) | 9 (22.0) | $< 0.0001$ |
| History of Pregnancy - n (%) | 4 (9.3) | 21 (51.2) | $< 0.0001$ |
| History of STI<sup>a</sup> - n (%) | 6 (14.0) | 5 (12.2) | 0.8113 |
| **Sexual Behaviors** |  |  |  |
| History of Sex for Money/Goods/Drugs - n (%) | 2 (4.7) | 5 (12.2) | 0.2594 |
| Age at First Sexual Encounter, years – mean (SD) | 17.2 (3.10) | 16.8 (2.49) | 0.5111 |
| New Partners in the past month – n (%) | 9 (22.0) | 12 (33.3) | 0.2632 |
| Number of Male Sexual Partners in the past month – median (IQR) | 1 (1 – 1) | 1 (1 – 2) | $0.0297$ |
| Number of Male Sexual Partners in the past 5 years – median (IQR) | 5 (2 – 11) | 3 (3 – 7.5) | 0.4759 |
| Condom use during vaginal sex in last month – n (%) |  |  | 0.8330 |
| Always | 6 (17.1) | 8 (22.9) |  |
| Sometimes | 10 (28.6) | 9 (25.7) |  |
| Never | 19 (54.3) | 18 (51.4) |  |
| History of substance use<sup>b</sup> – n (%) | 33 (76.7) | 16 (39.0) | $0.0005$ |
| **HPV Knowledge, HPV testing and Vaccination Practices** |  |  |  |
| Cumulative HPV Score, 29-item - mean (SD) | 18.9 (6.05) | 9.1 (8.82) | $< .0001$ |
| HPV Knowledge Score, 16 item – mean (SD) | 11.0 (3.97) | 5.9 (5.46) | $< .0001$ |
| HPV Testing Score, 6-item – mean (SD) | 3.1 (1.81) | 1.3 (1.78) | $< .0001$ |
| HPV Vaccination Score, 7-item – mean (SD) | 4.8 (1.44) | 1.9 (2.64) | $< .0001$ |
| History of Pap Smear – n (%) | 33 (76.7) | 36 (87.8) | 0.1567 |

Abbreviations. IQR, interquartile range; PrEP, pre-exposure prophylaxis; SD, standard deviation; STI, sexually transmitted infections
<sup>a</sup> Gonorrhea, chlamydia, syphilis, and Trichomonas
<sup>b</sup> Medical or recreational marijuana, cocaine, crack, heroin, methamphetamine, hallucinogens, club drugs, or any other illicit or recreational drugs

### HPV Knowledge, Screening, and Vaccination Practices

On the cumulative HPV score, participants in the vaccinated group reported a mean score of 18.9 (SD = 6.05) compared to 9.1 (SD = 8.82) for unvaccinated individuals (p < 0.0001). Mean HPV sub-scores on the 16-item HPV knowledge (11.0 vs 5.9, p <0.0001), 6-item HPV testing (3.1 v 1.3, p < 0.0001), and 7-item HPV vaccination (4.8 vs 1.9, p < 0.0001) scores remained significantly higher for the vaccinated individuals compared to non-vaccinated individuals. Out of the 84 individuals, only 66 (78.6%) women had ever heard of HPV and 58 (69.1%) had ever heard of HPV vaccination. Among the group of 66 women who were aware of the virus, 60 (90.9%) knew that HPV infection could be sexually transmitted, 50 (89%) knew that having many sexual partners increases the risk of getting HPV, and 54 (81.8%) knew that HPV causes cervical cancer. Within the group aware of HPV vaccination, 54 (93.1%) knew that it could prevent cervical cancer. Out of the 43 vaccinated women, 3 (7.0%) reported receiving one dose, 5 (11.6%) two doses, and 31 (72.1%) three doses; 4 women (2.3%) did not know the exact number of doses received. The mean age at first HPV vaccine dose was 15 (SD = 5.14) years, and 49 (58.3%) women reported a history of HPV screening/Pap smear at least once.

### Barriers and Motivators to HPV vaccination

Among unvaccinated women, barriers to HPV vaccination included low-risk perception (14.6%), healthcare barriers (29.3%), and vaccine hesitancy and personal beliefs (46.3%) with specific reasons shown in Table 3. The commonly reported motivators or reasons for willingness to receive the HPV vaccine among both vaccinated and unvaccinated women included protecting one’s health (67.8%), doctor recommendations (51.2%), and preventing the spread of HPV (51.2%) (Table 4). Significant group differences between vaccinated to unvaccinated women were seen in protecting one’s health (83.7% vs 51.2%; p = 0.0014), information on the risks associated with contracting HPV (44.2% vs 17.1%; p = 0.0072), doctor recommendations (69.8% vs 31.7%; p = 0.0005), preventing the spread of HPV (62.8% vs 39.0%; p = 0.0294), and discussion with parents or relatives (27.9% vs 0.0%; p = 0.0003).

**Table 3.** Barriers to HPV vaccination in unvaccinated participants (N = 41)

|  |  |
| --- | --- |
| **Low-Risk Perception - n (%)** | 6 (14.6) |
| “I am married/in a stable relationship” | 2 (2.4) |
| “I'm too old for the HPV vaccine” | 4 (4.8) |
| “Not sexually active” | 0 (0.0) |
| “Already have HPV, sexually active, or at-risk” | 0 (0.0) |
| **Healthcare Barriers - n (%)** | 12 (29.3) |
| “My doctor has never discussed it with me” | 11 (13.1) |
| “Cost/no private insurance” | 3 (3.6) |
| **Vaccine Hesitancy and Personal Beliefs - n (%)** | 19 (46.3) |
| “I've never really thought about it” | 10 (11.9) |
| “I don't know enough about it” | 6 (7.1) |
| “I'm not sure it's safe” | 4 (4.8) |
| “Potential health risks” | 2 (2.4) |
| “Potential side effects” | 3 (3.6) |
| “Product(s) haven't been around long enough” | 0 (0.0) |
| “I am still undecided” | 2 (2.4) |
| “I don't like needles” | 1 (1.2) |
| “I don't agree with vaccination” | 0 (0.0) |
| “Not aligned with my religious/cultural beliefs” | 0 (0.0) |
| “The HPV vaccine doesn't work” | 0 (0.0) |

**Table 4.** Motivators to HPV vaccination among vaccinate and unvaccinated participants (N = 84)

|  | Vaccinated<br>N = 43<br>(51.2%) | Unvaccinated<br>N = 41 (48.8%) | P-value |
| --- | --- | --- | --- |
| **Risk Perception and Vaccine Beliefs - n (%)** | 37 (86.1) | 23 (56.1) | **0.0024** |
| “Protecting my health” | 36 (83.7) | 21 (51.2) | **0.0014** |
| “Information on the risks associated with contracting HPV” | 19 (44.2) | 7 (17.1) | **0.0072** |
| “Information on the efficacy of the vaccine” | 11 (25.6) | 6 (14.6) | 0.2119 |
| “Information on the safety of the vaccine” | 6 (14.0) | 7 (17.1) | 0.6927 |
| “I know someone who has HPV” | 2 (4.7) | 4 (9.8) | 0.4274 |
| “I know someone who has had cervical cancer” | 1 (2.3) | 4 (9.8) | 0.1965 |
| **Healthcare-Related - n (%)** | **34 (79.1)** | **17 (41.5)** | **0.0004** |
| “A recommendation from my doctor” | 30 (69.8) | 13 (31.7) | **0.0005** |
| “Discussions with my doctor” | 16 (37.2) | 9 (22.0) | 0.1263 |
| “Vaccine was covered by health insurance” | 14 (32.6) | 7 (17.1) | 0.1014 |
| **Social - n (%)** | **31 (72.1)** | **16 (39.0)** | **0.0023** |
| “Preventing the spread of HPV” | 27 (62.8) | 16 (39.0) | **0.0294** |
| “Discussions with my parents or relatives” | 12 (27.9) | 0 (0.0) | **0.0003** |
| “One of my friends got vaccinated” | 2 (4.7) | 4 (9.8) | 0.4274 |
| “Mandatory at school” | 3 (7.0) | 3 (7.3) | 1.0000 |
| “Offered at school” | 1 (2.3) | 3 (7.3) | 0.3540 |

### Factors associated with HPV vaccination: Multivariate logistic regressions

Covariates included in the first multivariate logistic regression analysis were the cumulative HPV score, age, race, ethnicity, college education status, current contraceptive use, and number of sexual partners (Fig 1 & 2). This model had no violations in multicollinearity (CNI = 10.8) and had good fit (X^2^ = 8.9, p = 0.349). A one-year increase in age was associated with a 16% lower odds of HPV vaccination (aOR = 0.84, 95% CI [0.72, 0.99]; p = 0.035) while controlling for the other covariates in the model. Women who were using contraceptives had 8.36 (95% CI [1.41, 49.62]; p = 0.020) times the odds of HPV vaccination compared to women not on contraceptives. Race, ethnicity, college education status, and number of sexual partners were not significant predictors of HPV vaccination (Fig 1). HPV score (estimate = 0.1732, p = 0.0086) and HPV score^2^ (estimate = -0.0148, p = 0.0344) were significant predictors of HPV vaccination. The odds of HPV vaccination follow a quadratic relationship with diminishing odds of HPV vaccination after every one-point increase in HPV score. In fact, one-point increases in HPV score were significantly associated with higher odds of HPV vaccination until an HPV score of 16 (Fig 2).

**Fig 1.**
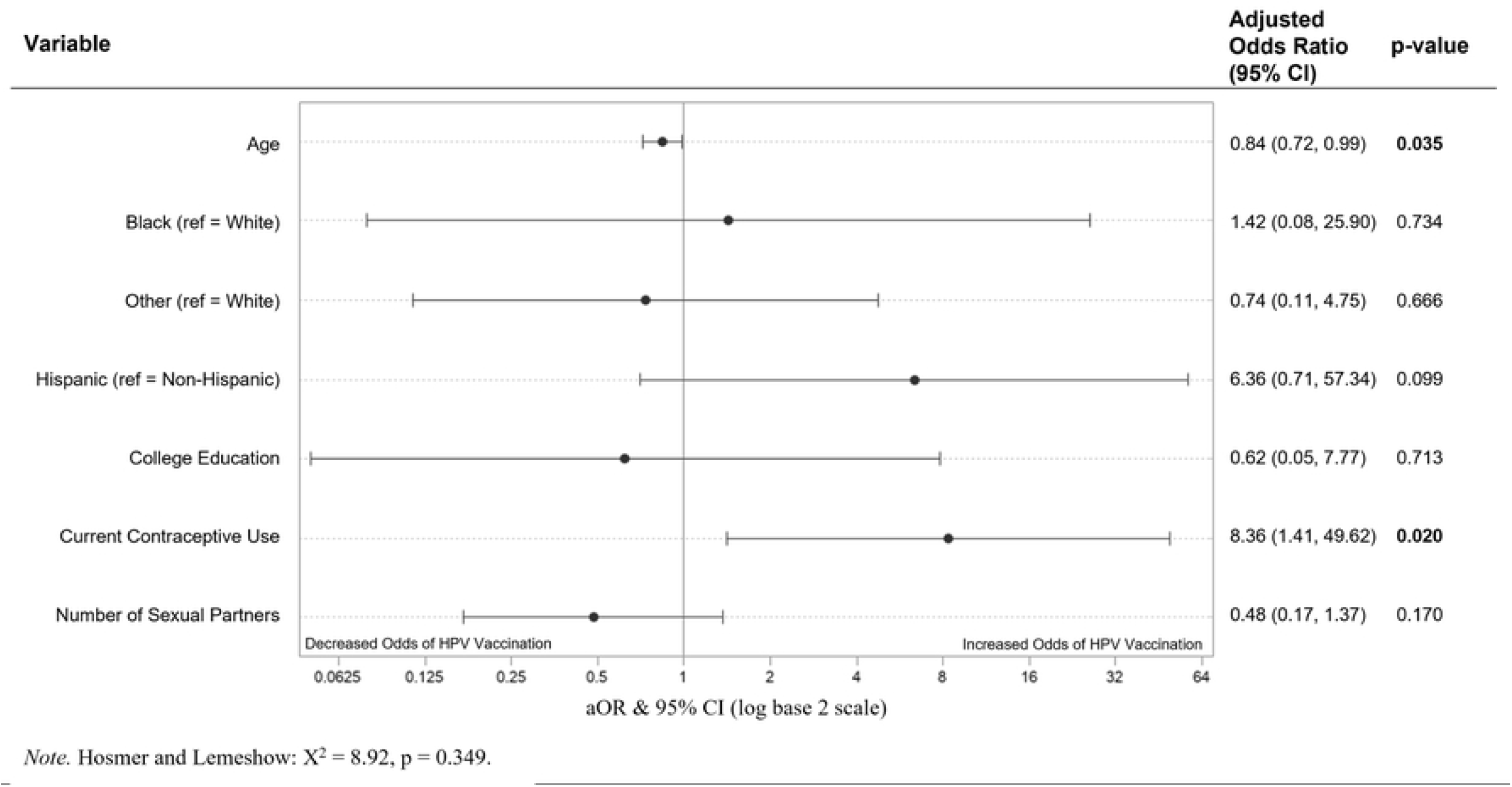
Predictors of HPV Vaccination in Multivariate Logistic Regression (n = 84). Predictors of HPV vaccination in a multivariable logistic regression model containing age, race, ethnicity, educational attainment, current contraceptive use, number of sexual partners, and HPV score. Adjusted Odd Ratios of adverse events were calculated and presented using a log base 2 scale in a Forest Plot. Null line is indicated for no predictor effects and bolded lines represent adjusted odds ratio with 95% confidence intervals. Bolded lines above and below the null line indicate increased or decreased odds of adverse events, respectively.

**Fig 2.**
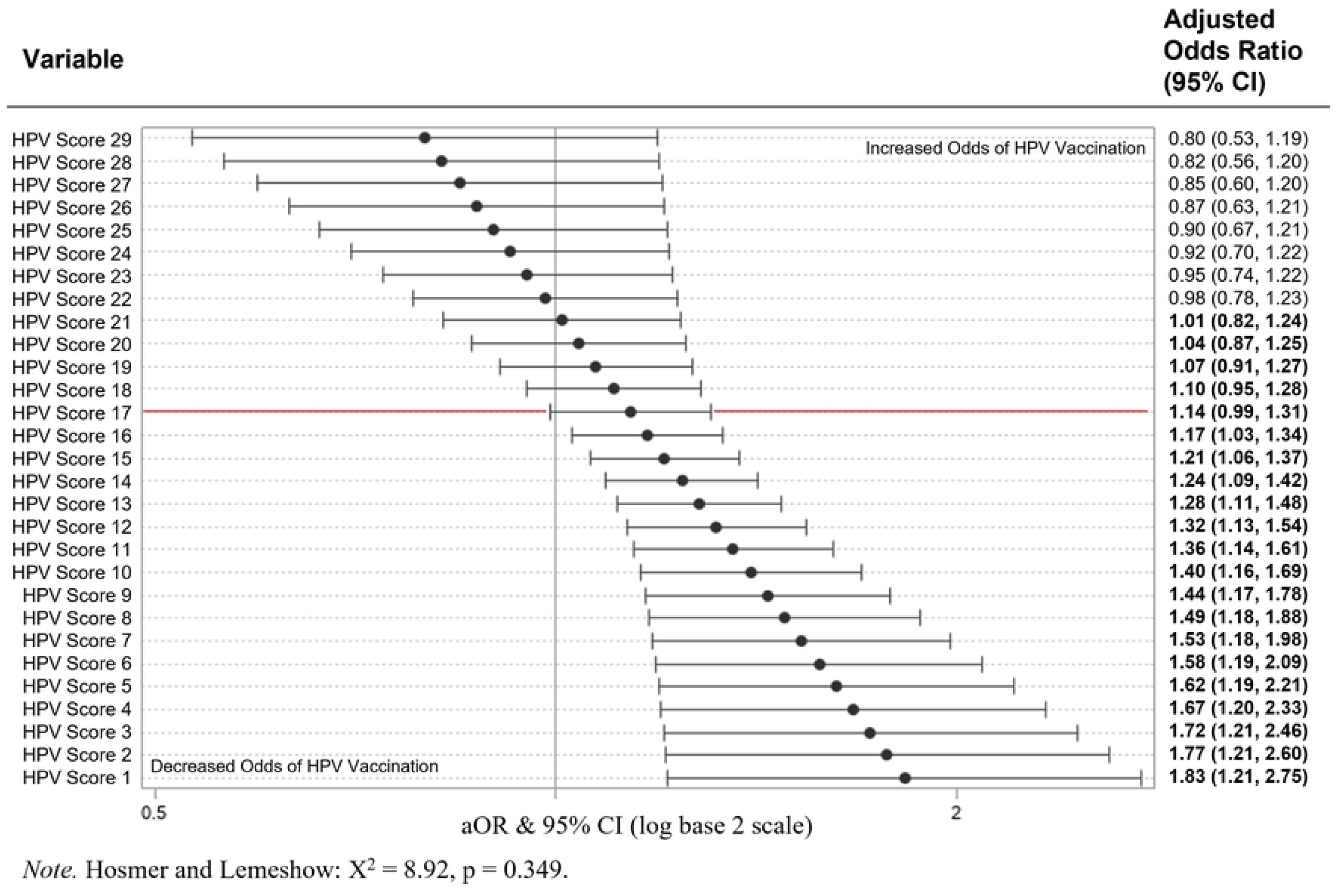
HPV Score as a Predictor of HPV Vaccination in Multivariate Logistic Regression (n = 84). HPV Score as a predictor of HPV vaccination in a multivariable logistic regression model containing age, race, ethnicity, educational attainment, current contraceptive use, number of sexual partners, and HPV score. HPV Score (p = 0.009) and higher order quadratic term of HPV Score (p = 0.034) are significant predictors of HPV vaccination. Single score changes beyond an HPV score of 16 are not associated with HPV vaccination. Adjusted Odd Ratios of adverse events were calculated and presented using a log base 2 scale in a Forest Plot. Null line is indicated for no predictor effects and bolded lines represent adjusted odds ratio with 95% confidence intervals. Bolded lines above and below the null line indicate increased or decreased odds of adverse events, respectively.

The second model selected evaluated vaccination motivators as predictors of HPV vaccination. Covariates fitted included collapsed categories of vaccination motivators: risk perception and vaccine beliefs, healthcare related, and social motivators (Table 4). The second model did not have any multicollinearity violations (CNI = 5.04) and had good fit (X^2^ = 3.88, p = 0.422).

Individuals who were motivated by healthcare had 3.03 (95% CI [1.02, 9.00]; p = 0.046) times the odds of HPV vaccination compared to individuals without healthcare-related motivators, while controlling for risk perception and vaccine beliefs, and social motivators (Fig 3). Risk perception and vaccine beliefs (aOR = 1.98, 95% CI [0.68, 5.76] ; p = 0.208) and social (aOR = 2.47, 95% CI [0.75, 8.07]; p = 0.135) motivators did not significantly predict HPV vaccination.

**Fig 3.**
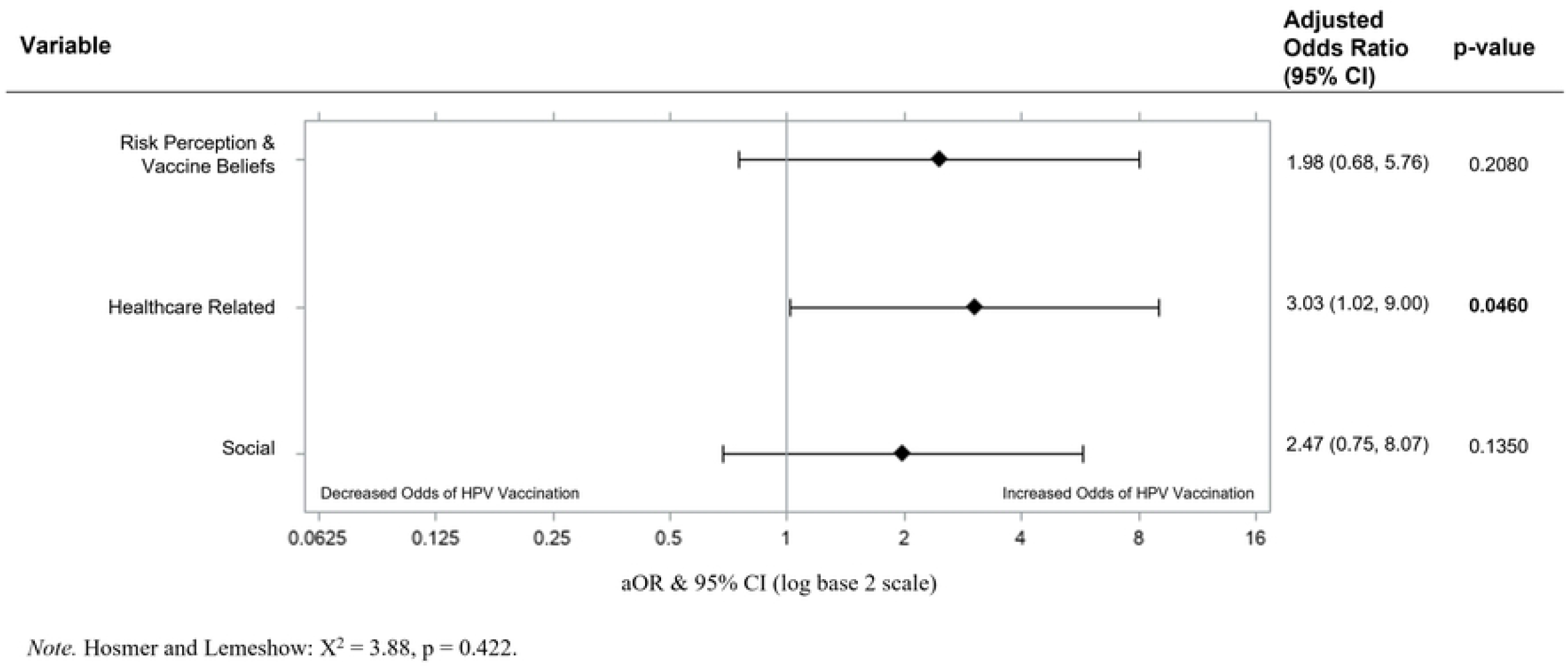
Motivators of HPV Vaccination in Multivariate Logistic Regression (n = 84). Motivators of HPV vaccination in a multivariable logistic regression model containing risk perception & vaccine beliefs, healthcare related, and social motivators. Adjusted Odd Ratios of adverse events were calculated and presented using a log base 2 scale in a Forest Plot. Null line is indicated for no predictor effects and bolded lines represent adjusted odds ratio with 95% confidence intervals. Bolded lines above and below the null line indicate increased or decreased odds of adverse events, respectively.

## DISCUSSION

This cross-sectional study evaluated HPV knowledge, screening, and vaccination practices as well as factors associated with vaccination among reproductive-age women living in Miami, Florida. It also evaluated barriers and motivations to HPV screening and vaccination. Findings suggest low vaccination uptake and screening practices amongst all the participants, and lower HPV knowledge was seen in the unvaccinated women compared to the vaccinated women. Our study also found that the cumulative HPV score, current contraceptive use, participant’s age, and healthcare-related motivators were significant predictors of HPV vaccination. Our results highlight the importance of increasing general HPV knowledge in women in a high-risk area particularly in those with little-to-no HPV knowledge, in order to increase vaccine uptake. To our knowledge, this is the first study in South Florida reporting on the use of a validated HPV score as a predictor of HPV vaccination, and to identify factors that can potentially be modified to increase HPV vaccine uptake.

Prior studies have shown that increased knowledge of HPV was correlated with vaccination status^12,23,24^. Similarly, our study found that cumulative HPV score was highly correlated with vaccination status. The mean score of the overall 29-item questionnaire for the vaccinated group was more than double that of the unvaccinated group. An important finding to note is that the likelihood of vaccination increased for every 1-point increase in HPV score until reaching a plateau at a score of 16/29. Above 16, a 1-point increase in HPV score was not a significant predictor of HPV vaccination. This suggests that overall vaccination rates would increase by improving basic knowledge of HPV in those with little-to-no knowledge, as opposed to improving knowledge in those who already have a foundational knowledge of HPV. Educational interventions should target populations with minimal HPV knowledge to improve vaccination rates.

Prior studies have suggested using routine reproductive health visits, such as contraceptive counseling visits, are important opportunities for providers to educates patients about HPV vaccination^25,26^. Our results are in alignment with this belief as there were more participants vaccinated among those who reported current contraception use. The strong correlation between contraceptive use and vaccination status could be related to multiple factors. Because contraception is provided by physicians, these participants presumably have frequent interaction with the healthcare system, increasing the likelihood that a provider will discuss HPV vaccination with them. Furthermore, these participants may have a higher degree of knowledge of sexual behaviors and risks, allowing them to see the benefit of the HPV vaccine. With regard to barrier contraception, a high number of participants who reported never using condoms during vaginal sex in the last month were unvaccinated. This serves as a reminder that there is still a subset of patients that can be counseled on HPV vaccines at their contraceptive health visits as well as HPV risk reduction through condom use.

The HPV vaccine became medically available in 2006, and in 2018 the FDA expanded the use of Gardasil 9, the current HPV vaccine, to include individuals ages 27 to 45. Prior to this date, the vaccine was only approved for ages 9 to 26.^27^ The difference in age distribution among vaccinated and unvaccinated participants seen in our study is likely a result of the historic vaccination guidelines and is congruent with the current literature.^28,29^ For the older participants in our study, HPV vaccination was less likely to have been recommended in their youth when they were more engaged with the health care system and followed routine immunization scheduling. In contrast, the younger participants were likely recommended HPV vaccination at pediatrician visits when they were adolescents. Currently the American College of Obstetrics and Gynecology and the Advisory Committee on Immunization Practices do not recommend catch-up vaccination for individuals age 27-45 but shared clinical decision making depending on the patient’s risk for new HPV infections.^30,31^ Our results suggest that there could be a subset of older reproductive age women, that could still benefit from HPV risk factor screening and counseling.

Prior research found that barriers to HPV vaccination vary depending on the participant population studied^32-35^. Of the barriers to HPV vaccination identified among unvaccinated women in our analysis, the majority were related to vaccine hesitancy and personal beliefs, and less due to low knowledge or healthcare access. These findings highlight the importance of promoting HPV vaccination, both within healthcare systems as well as throughout the community. Low-risk perception barriers were only reported by about 15% of unvaccinated women which could suggest a limited understanding of HPV risk factors.

Studies have also shown that provider recommendation is one of the strongest predictors of HPV vaccination^19^, and we see this in our study with 69.8% of vaccinated participants attesting to provider recommendation as a main reason for vaccination. Among the motivators for HPV vaccination, healthcare related motivators were found to increase odds of HPV vaccination by 300% when compared to individuals who did not identify these motivators. Based on this finding, increasing vaccination rates would be best accomplished through healthcare related routes, with a focus on provider recommendations.

A large limitation to this study is the sample size as it restricted the number of variables we could include in the multivariate analysis and thus models are subjected to uncontrolled confounders. Only 84/100 participants knew their vaccination status, limiting our sample but pointing to low HPV health literacy and health history knowledge in this population. Furthermore, this was a cross-sectional study which is limited by temporality. Ultimately, a cause-and-effect relationship cannot be determined with this study design. Our participant recruitment from prior study registries is another limitation to the generalizability of our results as it potentially introduces sampling bias. Providing surveys only in English presents another limitation in the ethnically diverse community of Miami.

Regardless of the limitations, this is an important study as it was conducted in a Southern US city with a high incidence of STIs and low HPV vaccination coverage. Despite an English-only survey, our participant population was racially and ethnically diverse. A strong component of this study is the use of validated questionnaires to assess HPV knowledge, screening, and vaccination practices as well as barriers and motivators. As the answers to these questions were found to be significantly different between the vaccinated and unvaccinated groups, it is important that the validity of these questions was confirmed. Moreover, the study design was strengthened by providing participants the option to complete the survey through a secure web-based system, likely reducing the risk of inaccurate responses that could occur if participants were concerned about privacy.

Overall, this study was successful in evaluating HPV knowledge, screening, and vaccination practices among women of reproductive age living in Miami. Future studies should recruit larger sample sizes and include surveys in Spanish, Haitian Creole and other languages in order to gain a better understanding of the racial and ethnic disparities related to HPV vaccination. The clear relationship between HPV knowledge and vaccination status further emphasizes the need to educate communities about HPV to reduce the spread of the disease and ultimately reduce the rates of HPV-related cancers.

## Data Availability

All relevant data are within the manuscript.

## ACKNOWLEDGEMENTS

We thank the members of the Miami CFAR (Center for AIDS research) for their support and collaboration in the study.

## Abbreviations

(MLR): Multivariable logistic regressions
(UM): University of Miami
(HPV): Human papillomavirus

## SUPPORTIVE INFORMATION

None

